# Pneumocytes are distinguished by highly elevated expression of the ER stress biomarker GRP78, a co-receptor for SARS-CoV-2, in COVID-19 autopsies

**DOI:** 10.1101/2021.06.17.21259098

**Authors:** Andrii Puzyrenko, Elizabeth R. Jacobs, Yunguang Sun, Juan Felix, Yuri Sheinin, Linna Ge, Shuping Lai, Qiang Dai, Rahul Nanchal, Paula North, Pippa Simpson, Hallgeir Rui, Ivor J. Benjamin

## Abstract

Vaccinations are widely credited with reducing death rates from COVID-19 but the underlying host-viral mechanisms/interactions for morbidity and mortality of SARS-CoV-2 infection remain poorly understood. Acute respiratory distress syndrome (ARDS) describes the severe lung injury, which is pathologically associated with alveolar damage, inflammation, non-cardiogenic edema, and hyaline membrane formation. Because proteostatic pathways play central roles in cellular protection, immune modulation, protein degradation and tissue repair, we examined the pathological features for the unfolded protein response (UPR) using the surrogate biomarker glucose regulated protein 78 (GRP78) and co-receptor for SARS-CoV-2. At autopsy, immunostaining of COVID-19 lungs showed highly elevated expression of GRP78 in both pneumocytes and macrophages compared to non-COVID control lungs. GRP78 expression was detected in both SARS-CoV-2 infected and un-infected pneumocytes as determined by multiplexed immunostaining for nucleocapsid protein. In macrophages, immunohistochemical staining for GRP78 from deceased COVID-19 patients was increased but overlapped with GRP78 expression taken from surgical resections of non-COVID-19 controls. In contrast, the robust *in situ* GRP78 immunostaining of pneumocytes from COVID-19 autopsies exhibited no overlap and was independent of age, race/ethnicity and gender compared with non-COVID-19 controls. Our findings bring new insights for stress-response pathways involving the proteostatic network implicated for host resilience and suggest that targeting of GRP78 expression might afford an alternative therapeutic strategy to modulate host-viral interactions during SARS-CoV-2 infections.

## Introduction

The coronavirus disease-19 (COVID-19) pandemic has afflicted over 33 million people, causing approximately 5 million hospitalizations and nearly 600,000 deaths in the United States alone (>4 million deaths worldwide). Understanding mechanisms through which the severe acute respiratory disease coronavirus 2 (SARS-CoV-2) infection promotes major pathophysiological consequences of COVID-19 is key to improving strategies for monitoring, mitigation and/or treatment. Host stress-response pathways orchestrate a cascade of cellular events as the first line of defense to preserve cellular homeostasis by maintaining proper protein folding, i.e. ‘**proteostasis**’.^1,2^ For example, the endoplasmic reticulum (ER)-localized glucose-regulated protein 78 (GRP78/HSPA5), a molecular chaperone and mediator of the unfolded protein response (UPR), plays a key role involving the proteostatic network/machinery during ER stress^3,4^ and has also has been implicated in host-viral interactions. While the Spike protein of SARS-CoV-2 binds to the ACE2 receptor in humans, host tropism is also determined by GRP78/HSPA5, which acts as a coreceptor on the cell surface and promotes viral entry for the SARS-CoV, middle east respiratory syndrome coronavirus (MERS-CoV), bat coronavirus^5^, and has been predicted to bind SARS-CoV-2 Spike protein/ACE2 complexes.^6,7^ Because viral genomes lack heat shock proteins (HSPs), invasion of the single-strand RNA SARS-CoV-2 virus is obligatorily coupled with, and dependent upon, recruitment of the host’s protein synthesis machinery to maintain folding of viral proteins. Competition for the host’s HSP/GRP78 machinery creates existential threats for both host and virus, invoking complex cell fate decisions resulting in death or survival. Recent evidence for the appearance of plasma UPR biomarkers with COVID-19 suggests early tissue damage associated with the hyperinflammatory state. However, insight about clinical progression to severe disease^8^, critical for timely management of COVID-19 patients^9^ is lacking. We hypothesized that host-viral interactions of SARS-CoV-2 are conceivably accompanied by defects either in the components of the proteostasis machinery, dysregulation of normal proteostatic signaling or both. GRP78/HSPA5 is expressed in the normal respiratory airway epithelial cells and to a lesser extent in normal alveolar pneumocytes, and can facilitate pathogenic viral entry into the cells^10^. Elevated levels of circulating grp78 mRNA have been reported in COVID-19 patients^10^. We therefore focused on the expression of GRP78 in lungs of patients suffering COVID-19 deaths with respiratory failure.

## Materials and Methods

### Materials

A monoclonal rabbit antibody to GRP78 (BiP (C50B12) Rabbit mAb; Cell Signaling, Danvers, MA; Cat#3177; 1:200) and Envision Plus polymer (Agilent/DAKO) were used with magenta chromogen for visualization. Immunohistochemistry for SARS-CoV-2 Nucleocapsid protein was performed using an affinity-purified polyclonal rabbit antibody (affinity-purified rabbit IgG; ProSci, Poway, CA; Cat# 9099; 0.04 µg/ml). Multiplex immunofluorescence staining of GRP78 was visualized using TSA Plus Cyanine 3 System, Perkin Elmer, Waltham, MA; Cat# NEL744B001KT). SARS-CoV-2 Nucleocapsid was visualized using TSA Plus Cyanine 5 System (Perkin Elmer Cat# NEL745001KT). Macrophages/monocytes were identified using CD68 (CD68 Monoclonal Antibody KP1, Thermo Fisher Scientific, Grand Island, NY; Cat# 14-0688-82) and visualized using TSA Plus Cyanine 7 System (Perkin Elmer, Cat# CUSM03644000EA) following peroxidase blocking reagent (hydrogen peroxide solution, Sigma Aldrich, St. Louis, MO; Cat# H1009). Antibody stripping was accomplished using the Omnis (Dako/Agilent) low (CD68 and SARS-CoV-2 Nucleocapsid) or high (GRP78) pH antigen retrieval protocol. Pneumocytes were identified using anti-pan-cytokeratin antibody (mouse monoclonal AE1/AE3 blend, Cat# M3515, Agilent) followed by Alexa-Fluor-488-labeled secondary antibody (Cat# A11029, Thermo Fisher Scientific).

### Immunohistochemistry

Immunohistochemistry was performed on deparaffinized sections of lungs from either COVID-19 decedents or from patients with non-COVID-19 conditions using an Omnis autostainer (Agilent/DAKO, Santa Clara, CA). A monoclonal rabbit antibody to GRP78 (BiP; C50B12; Cell Signaling, Danvers, MA; Cat#3177; 1:200 dilution) and Envision Plus polymer (Agilent/DAKO) were used with magenta chromogen for bright field visualization. For fluorescence immunohistochemistry, SARS-CoV-2 Nucleocapsid protein was performed using an affinity-purified polyclonal rabbit antibody (ProSci, Poway, CA; Cat# 9099) in a cross-validated protocol as previously described^11^. Multiplex immunofluorescence staining of GRP78, SARS-CoV-2 Nucleocapsid, and CD68 was performed following incubation with peroxidase blocking reagent (hydrogen peroxide solution, Sigma Aldrich, St. Louis, MO; Cat# H1009) using iterative incubation with a) primary antibody, b) secondary HRP-conjugated antibody, c) HRP-mediated deposition of fluorescent-tyramide, d) antibody stripping using the Omnis low pH (pH 6; CD68 and SARS-CoV-2 Nucleocapsid) or high pH (pH 9; GRP78) antigen retrieval protocols, repeated for each of the three primary antibodies. Specifically, immunofluorescence staining of GRP78 (C50B12; Cell Signaling, Danvers, MA; Cat#3177; 1:800 dilution for IF) was visualized using the TSA Plus Cyanine-3 System, Perkin Elmer, Waltham, MA; Cat# NEL744B001KT); SARS-CoV-2 Nucleocapsid (ProSci, Poway, CA; Cat# 9099; 0.01 µg/ml) was visualized using the TSA Plus Cyanine-5 System (Perkin Elmer Cat# NEL745001KT); macrophages were identified using CD68 (CD68 Monoclonal Antibody KP1, Thermo Fisher Scientific, Grand Island, NY; Cat# 14-0688-82; 1:10,000 dilution) and visualized using the TSA Plus Cyanine-7 System (Perkin Elmer, Cat# CUSM03644000EA). Iterative primary antibody stripping was accomplished using the Omnis (Dako/Agilent) low (pH 6; CD68 and SARS-CoV-2 Nucleocapsid) or high (pH 9; GRP78) antigen retrieval protocols. Pneumocytes were identified using anti-pan-cytokeratin antibody (mouse monoclonal AE1/AE3 blend, Cat# M3515, Agilent) followed by Alexa-Fluor-488-labeled secondary antibody (Cat# A11029, Thermo Fisher Scientific) and final counterstaining with DAPI. After coverslipping, chromogen IHC stained slides were scanned using bright field mode of Pannoramic 250 (3DHISTECH Ltd., Budapest, Hungary) and images were generated using Caseviewer, while multicolor immunofluorescence-stained slides were scanned on a Vectra Polaris (Akoya Biosciences) and images generated from PhenoCharts (Akoya Biosciences, Marlborough, MA).

Lung sections analyzed were taken from the most prominent areas of change on gross examination as determined by a pathologist from the COVID-19 positive group.

Sections from non-COVID-19 control specimens were selected from grossly unremarkable lung tissue. Each lung section of every case was evaluated microscopically in entirety at low and high magnifications by pathologists. Representative images showing the dominant histological features were taken at different magnifications (20x, 200x and 400x) from each case in both the COVID-19 and non-COVID-19 control groups for scoring.

### Scoring of GRP78 expression

GRP78 expression was evaluated in pneumocytes and macrophages by pathologists. The GRP78 staining intensity of either cell type was awarded the following 4 grades according to the staining intensity of magenta color as follows: 0 - no staining, 1 - weak, 2 - moderate, and 3 - strong. The proportion of either pneumocytes or macrophages of each tissue that were positive for GRP78 were also graded as follows: 1 - less than 5%; 2 - 5%-24%, 3 - 25%-49%, 4 - 50%-74%, and 5 - 75%-100%. The sum of the GRP78 intensity grade and positivity grade for each cell type generated an overall GRP78 score that was represented by a 4 step scale as follows: grades 1-2 - negative (−); grades 3-4 - weak positive (+); grades 5-6 - moderate positive (++); and grades 7-8 - strong positive (+++).

### Statistical methods

Continuous variables are summarized as median (minimum, maximum) and groups are compared using exact Mann Whitney or Kruskal Wallis tests. Categorical variables are summarized as number (%) and compared using exact Fisher tests. p values are unadjusted and any P< 0.05 is denoted as significant. Software used was Statistical Package for Social Sciences v26.

## Results

Autopsy tissue from 12 patients diagnosed with COVID-19 (based on PCR tests and consistent CT chest, lab tests, and clinical exam) who were admitted through the emergency department of the Froedtert & the Medical College of Wisconsin regional hospital and died from SARS-CoV-2 pneumonia were included in the study. Archival non-COVID-19 control lung tissues were from resections (N=25; 14 cases of malignancy; samples secured from lung remote from that involved with tumor; and 11 cases showing inflammation/infection). For the control lung tissues with remote neoplasm, lungs were resected most commonly for either adenocarcinoma (n=8) or squamous cell carcinoma (n=4). For control lung tissues without malignancy, inflammation either with or without granulomatous changes was the most frequent pathological diagnosis (n=11), followed by either interstitial lung disease or fibrosis (n=4). Pathological evidence of emphysema was evident in tissue of 3 cases with cancer and 4 cases with inflammatory changes. All samples from COVID-19 autopsy patients demonstrated diffuse alveolar damage throughout the lungs.

Demographic information of the patient populations is shown in Table 1. Age and gender distribution of COVID-19 patients was similar to controls, including those with either cancer or inflammatory diseases. Of the COVID-19 patients, 67% were female, while 45% and 57% of the non-COVID-19/no cancer cases and non-COVID-19/remote cancer was female, respectively (p=0.58). Median (range) ages were 69 (28-91) for COVID-19 cases; 60 (22-70) for non-COVID-19/no cancer cases and 62.5 (56-85) for non-COVID-19/remote cancer cases (p=0.19). The proportion of Black cases was higher among COVID-19 cohort (7 Blacks, 5 Whites) compared to non-COVID-19 controls (1 Blacks, 11 Whites; p<0.001). These data are consistent with the reported greater burden of COVID-19 in Black Americans.^12^

**Table 1.** Shows age, gender, ethnicity/race of patients from COVID-19 and non-COVID-19 groups, as well as p values comparing demographic data between the groups. Representation of Black patients was greater in the Covid-19 relative to other groups. On the other hand, age distribution and gender were not different between donors from control and COVID-19 groups.

| Table 1 | COVID-19 | Controls | p value |
| --- | --- | --- | --- |
| Age median (min-max) | 69 (28-91) | 60 (22-85) | 0.102 |
| Gender (% female) | 0.67 | 0.52 | 0.49 |
| Black | 0.58 | 0.04 | 0.001 |
| Hispanic | 0.17 | 0 | 0.09 |
| White | 0.25 | 0.92 | 0.48 |

To determine the status of GRP78 protein expression in lungs of COVID-19 patients, we performed immunohistochemistry of GRP78. Figure 1 shows representative images of lungs from COVID-19 and non-COVID-19 patients stained for GRP78 protein expression by immunohistochemistry. Both intensity and distribution for GRP78 expression (magenta staining) were easily visible and more abundant in samples from COVID-19 patients compared with control patients. Parallel multiplexed co-immunostaining verified abundant GRP78 expression in a large proportion of both CD68^+^ macrophages and pan-cytokeratin positive pneumocytes in lung tissue from a patient who died during the active viral infection phase (Figure 2a,b). GRP78 was also expressed within actively infected pneumocytes, as visualized by co-expression of SARS-CoV-2 Nucleocapsid protein (Figure 2c-e).

**Figure 1.**
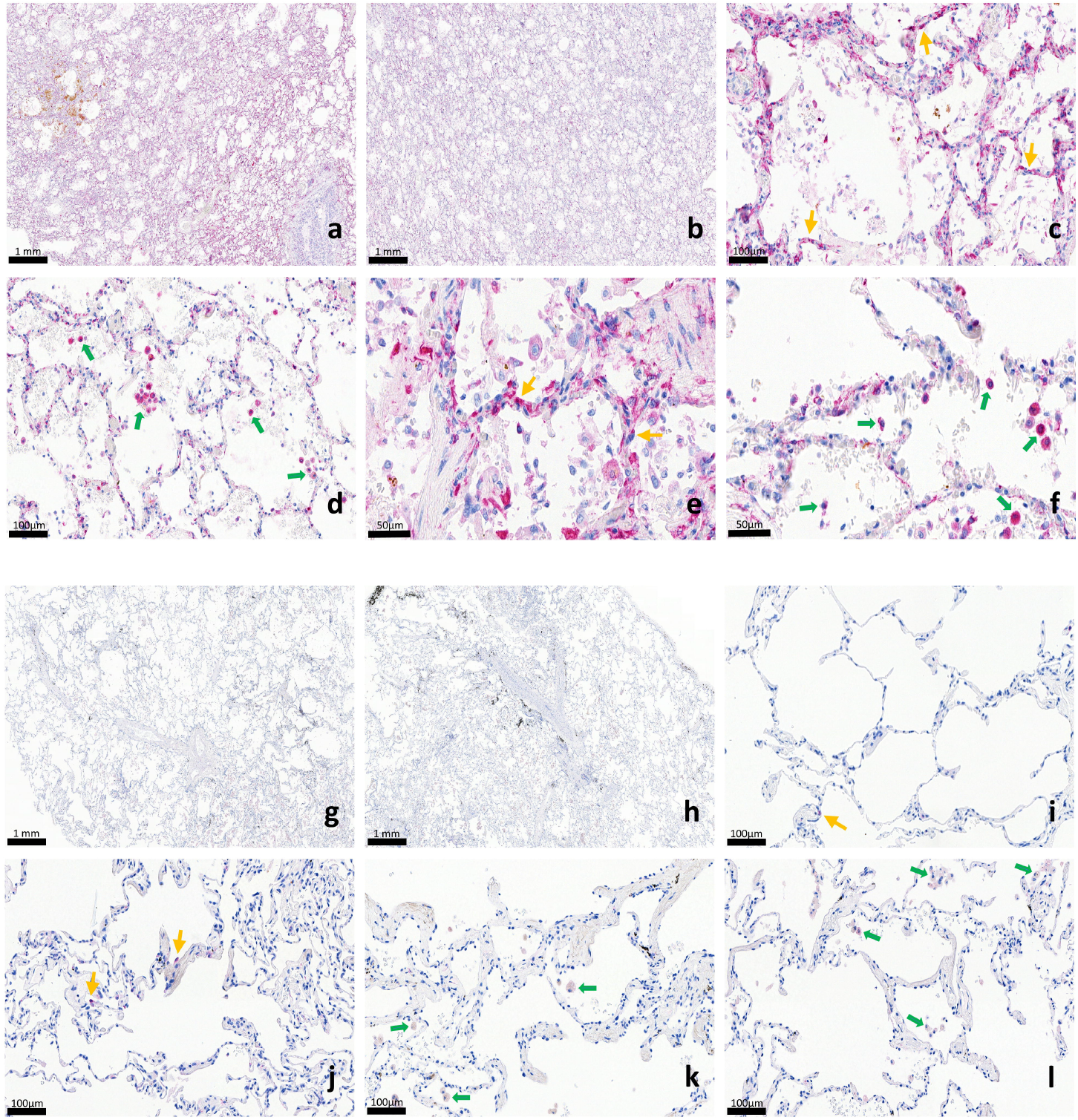
GRP78 protein expression among the lung sections from patients with COVID-19 (upper panels) and non-COVID-19 controls (bottom panels; a, b, g, h at 20x; c, d, i, k, l at 200 X and e,f at 400x). Yellow arrows indicate pneumocytes and green arrows identify macrophages. ***a, b)*** Representative overview histology images showing intense GRP78 expression in COVID-19 lung; ***c-f)*** Moderate to strong positive pneumocytes and macrophages on higher magnification in COVID-19 lung; ***g, h)*** Overview histology showing lung from non-COVID-19 control group; ***i-l)*** Rare positive pneumocytes and weak to moderate positive macrophages on higher magnification in lung from non-COVID-19 controls.

**Figure 2.**
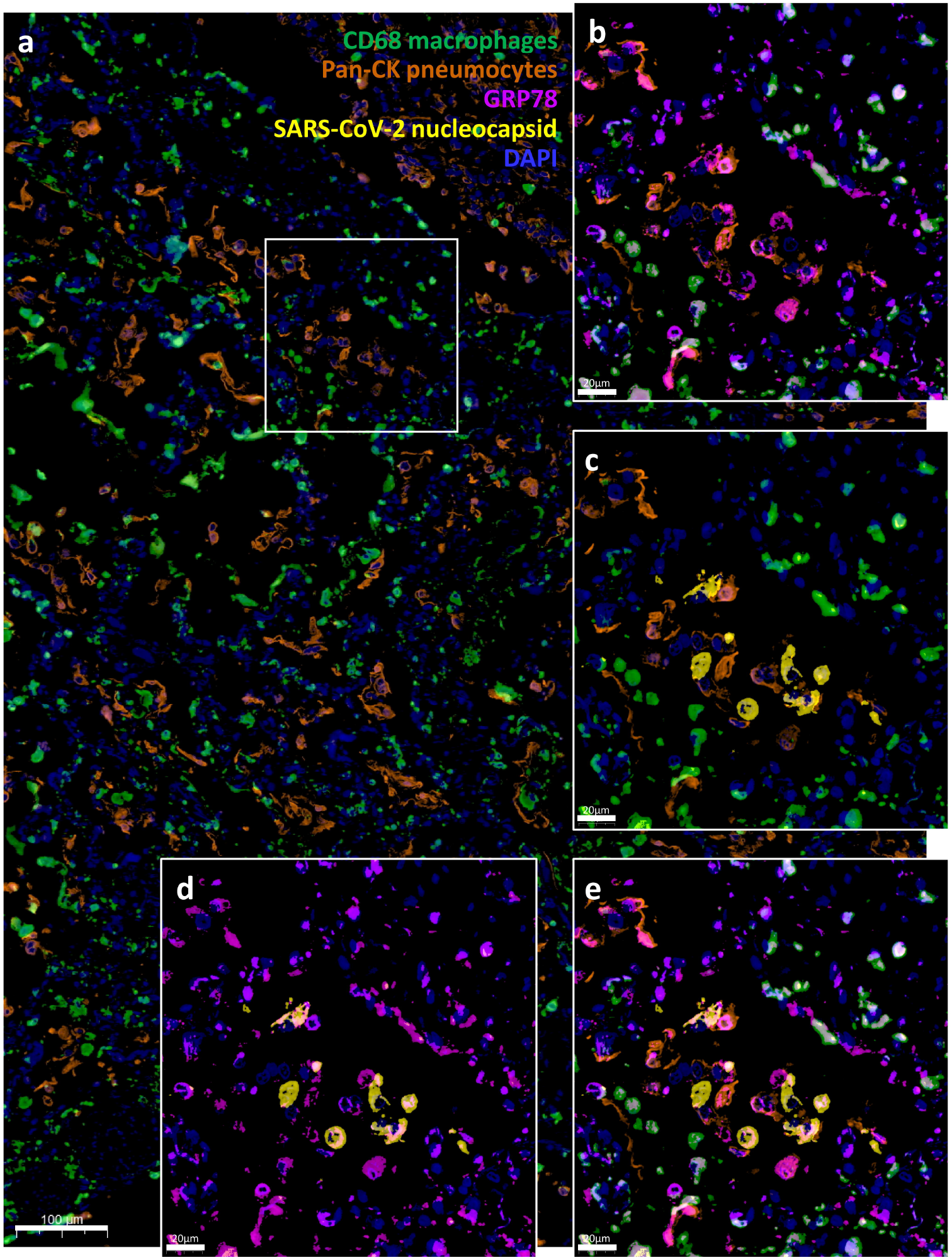
Multiplexed immunostaining of COVID-19 lung showing GRP78 expression in macrophages and pneumocytes. ***a)*** Overview histology showing pneumocytes (pan-cytokeratin; brown) and macrophages (CD68; green) with nuclei visualized by DAPI (blue); ***b)*** Higher power frame from a) with pneumocytes (brown) and macrophages (green) and merged with GRP78 staining (purple); ***c)*** same frame with pneumocytes (brown) and macrophages (green) and merged with SARS-CoV-2 nucleocapsid staining (yellow); ***d)*** same frame with GRP78 (purple) merged with SARS-CoV-2 nucleocapsid staining (yellow), and ***e)*** all five colors merged.

Table 2 shows the distribution of GRP78 staining intensity grades and percent GRP78-positivity grades for pneumocytes and macrophages on a case-by-case basis, and the two grades were added to yield a composite intensity plus positivity GRP78 score for each of the two cell types. Moderate to high GRP78 staining scores in the pneumocytes in lungs from COVID-19 patients were markedly higher than those of pneumocytes in non-COVID-19 control lungs (p<0.001). Similarly, strong positive scores of GRP78 in the macrophages were higher in the COVID-19 group compared to non-COVID-19 controls (P<0.001). Interestingly, the higher score for GRP78 expression in macrophages from COVID-19 patients was attributable to higher intensity of staining rather than the percentage of GRP78-positive cells. The subgroup of non-COVID-19 control with resections for cancer exhibited similar differences between pneumocyte scores relative to that of COVID-19 patients (COVID-19: median 8 (n=6-8); Cancer median 3 (n=24); p<0.001). When the analysis excluded data from Black cases, similar differences in scoring for pneumocyte and macrophage GRP78 expression between COVID-19 and control cases persisted (14.6 and 11.5 for COVID-19 patients respectively; p<0.001).

**Table 2.** Show scores for GRP78 intensity and percent positive cells on a patient by patient basis. Data are separated into those from patients with COVID-19, those with largely inflammatory process and those from lungs removed for cancer, tissue remote from the site of cancer. Median values and ranges are provided for each endpoint, as well as p values comparing values from groups of patients. Both the intensity of staining (on a scale of 1-3) and percent of pneumocytes positive (less than 5% - 1; 5%-24% - 2, 25%-49% - 3, 50%-74% - 4, 75%-100% - 5) are increased in samples from COVID-19 patients relative to that of samples from controls, as are the scores which represent a composite of these values. This observation also holds for values of patients with lung resection for remote cancer. The intensity of staining in macrophages was similar in COVID-19 and control patients, whereas the percentage of macrophages and, thereby the overall score for GRP78 staining, were higher in COVID-19 compared to that of other samples.

| Patient | 1 | 2 | 3 | 4 | 5 | 6 | 7 | 8 | 9 | 10 | 11 | 12 | 13 | 14 | 15 | 16 | 17 | 18 | 19 | 20 | 21 | 22 | 23 | 24 | 25 | 26 | 27 | 28 | 29 | 30 | 31 | 32 | 33 | 34 | 35 | 36 | 37 |  |
| --- | --- | --- | --- | --- | --- | --- | --- | --- | --- | --- | --- | --- | --- | --- | --- | --- | --- | --- | --- | --- | --- | --- | --- | --- | --- | --- | --- | --- | --- | --- | --- | --- | --- | --- | --- | --- | --- | --- |
| Diagnosis | COVID-19 |  |  |  |  |  |  |  |  |  |  |  | Control- inflammatory processes |  |  |  |  |  |  |  |  |  | Control- remote from cancer |  |  |  |  |  |  |  |  |  |  |  |  |  |  |  |
| Pneumocyte<br>GRP78 intensity | 3 | 3 | 3 | 3 | 3 | 2 | 3 | 3 | 3 | 3 | 3 | 3 | 2 | 2 | 2 | 2 | 2 | 2 | 1 | 1 | 2 | 1 | 2 | 2 | 2 | 2 | 2 | 2 | 2 | 1 | 2 | 2 | 2 | 2 | 2 | 2 | 2 |  |
| Pneumocytes pct<br>pos | 5 | 5 | 4 | 3 | 3 | 5 | 4 | 5 | 5 | 5 | 5 | 5 | 1 | 1 | 1 | 1 | 1 | 1 | 2 | 2 | 1 | 1 | 2 | 2 | 1 | 1 | 1 | 1 | 1 | 1 | 1 | 1 | 1 | 1 | 1 | 1 | 1 |  |
| Scale Pneumocytes | 8 | 8 | 7 | 6 | 6 | 7 | 7 | 8 | 8 | 8 | 8 | 8 | 3 | 3 | 3 | 3 | 3 | 3 | 3 | 3 | 3 | 2 | 4 | 4 | 4 | 3 | 3 | 3 | 3 | 3 | 2 | 3 | 3 | 3 | 3 | 3 | 3 |  |
| Macrophage<br>GRP78 intensity | 2 | 3 | 3 | 3 | 3 | 2 | 3 | 3 | 3 | 3 | 3 | 3 | 1 | 1 | 1 | 1 | 1 | 1 | 1 | 1 | 1 | 1 | 2 | 1 | 1 | 1 | 1 | 1 | 1 | 2 | 1 | 1 | 1 | 1 | 1 | 1 | 1 |  |
| Macrophage pct<br>pos | 5 | 5 | 4 | 3 | 3 | 5 | 4 | 5 | 5 | 5 | 5 | 5 | 5 | 4 | 5 | 3 | 3 | 4 | 5 | 5 | 4 | 4 | 4 | 5 | 4 | 4 | 4 | 5 | 5 | 5 | 3 | 4 | 5 | 5 | 4 | 5 | 4 |  |
| Scale Macrophages | 7 | 8 | 7 | 6 | 6 | 7 | 7 | 8 | 8 | 8 | 8 | 8 | 6 | 5 | 6 | 4 | 4 | 5 | 6 | 6 | 5 | 5 | 6 | 6 | 5 | 5 | 5 | 6 | 6 | 7 | 4 | 5 | 6 | 6 | 5 | 6 | 5 |  |
| COVID-19 |  |  |  |  |  |  |  |  |  |  |  |  | Control |  |  |  |  |  |  |  |  |  | Control- remote from cancer |  |  |  |  |  |  |  |  |  |  |  |  |  |  | p value COVID-19 vs all controls |
| Median (range)<br>pneumo intensity | 3(2-3) |  |  |  |  |  |  |  |  |  |  |  | 2(1-2) |  |  |  |  |  |  |  |  |  | 2(1-2) |  |  |  |  |  |  |  |  |  |  |  |  |  |  | <0.001 |
| Median (range)<br>pneumo pct pos | 5(3-5) |  |  |  |  |  |  |  |  |  |  |  | 1(1-2) |  |  |  |  |  |  |  |  |  | 1(1-2) |  |  |  |  |  |  |  |  |  |  |  |  |  |  | <0.001 |
| Median (range)<br>pneumo scale | 8(6-8) |  |  |  |  |  |  |  |  |  |  |  | 3(2-4) |  |  |  |  |  |  |  |  |  | 3(2-4) |  |  |  |  |  |  |  |  |  |  |  |  |  |  | <0.001 |
| Median (range)<br>macrophage<br>intensity | 3(2-3) |  |  |  |  |  |  |  |  |  |  |  | 1(1-2) |  |  |  |  |  |  |  |  |  | 1(1-2) |  |  |  |  |  |  |  |  |  |  |  |  |  |  | <0.001 |
| Median (range)<br>macrophage pct<br>pos | 5(3-5) |  |  |  |  |  |  |  |  |  |  |  | 4(3-5) |  |  |  |  |  |  |  |  |  | 4(3-5) |  |  |  |  |  |  |  |  |  |  |  |  |  |  | 0.28 |
| Median (range)<br>macrophage scale | 7.5(6-8) |  |  |  |  |  |  |  |  |  |  |  | 5(4-6) |  |  |  |  |  |  |  |  |  | 5(4-7) |  |  |  |  |  |  |  |  |  |  |  |  |  |  | <0.001 |

Figure 3 shows the distribution of GRP78 scores across all age ranges and stratified by COVID-19 versus non-COVID-19 controls. In pneumocytes (Figure 3a), there was a clear separation of GRP78 scores across all age groups with no overlap of patients with COVID-19 and other groups. Similarly, the same trend in the separation can be seen for GRP78 immunostaining staining in macrophages (Figure 3b), but unlike the data from pneumocytes, there was partial overlap in GRP78 scores in COVID-19 and non-COVID-19 controls.

**Figure 3.**
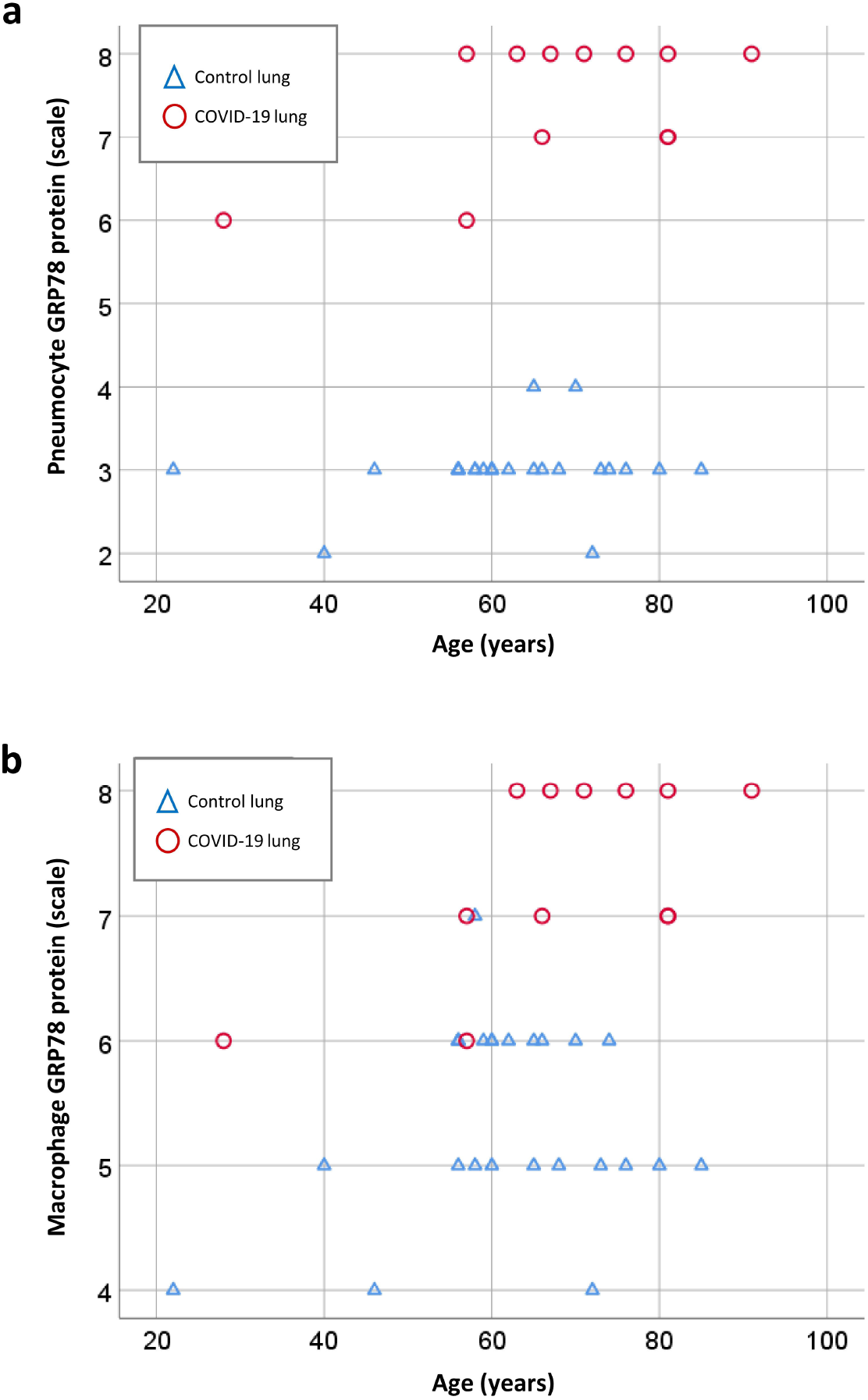
3a shows the scoring of pneumocytes for GRP78 staining as a function of diagnosis (COVID-19 or control states) and stratified by age. There is a clear separation of scores for GRP78 expression that is sustained across all age ranges studied. Figure 3b shows the same data for macrophages. Scores for COVID-19 are greater than those for non-COVID-19 controls, but there is overlap between the scores for group, unlike that of data for pneumocytes.

## Discussion

Our analyses show substantially increased expression of the stress protein GRP78 and SARS-CoV-2 co-receptor in pneumocytes and lung macrophages of patients who died while hospitalized for COVID-19 relative to those of non-COVID-19 control lungs. These observations may provide insights into the source of circulating GRP78 mRNA and protein in patients with SARS-COV-2-positive pneumonia that has been observed relative to individuals with SARS-COV-2-negative pneumonia or a healthy group^13^.

While GRP78 serves key cytoprotective and homeostatic effects in cells under some conditions, including viral infection and associated inflammatory states, GRP78 is also considered a co-receptor with Angiotensin-converting enzyme 2 (ACE2) for SARS-CoV-2. Our observation in COVID-19 pneumonia lungs, that elevated GRP78 expression levels were detected within both SARS-CoV-2 infected (nucleocapsid protein-positive) and un-infected (nucleocapsid protein-negative) pneumocytes as determined by multiplexed immunostaining, raises the possibility of a SARS-CoV-2 virus-GRP78 feed forward-loop that facilitates disease progression. In support of this notion, a humanized monoclonal antibody to GRP78 was recently shown to not only reduce cell surface ACE2 but to decrease SARS-CoV-2 entry and infection^14^. ACE2 expression is reported to increase with age in patients requiring mechanical ventilation^15^, raising the additional possibility that mechanical ventilation may contribute to the observed increased GRP78 levels in pneumocytes via enhanced ACE2 in the autopsied COVID-19 lungs. However, age-related increase in ACE2 expression from mechanical ventilation alone cannot account for our observations because increased GRP78 scores in pneumocytes and macrophages were observed across all age groups with COVID-19, without a relationship to age. Moreover, four of the 12 COVID-19 patients analyzed were not treated with mechanical ventilation; and the median GRP78 scores for pneumocytes and macrophages were not different from those who did receive mechanical ventilation (8 vs. 8 for pneumocytes; 7 vs. 8 for macrophages). Further work will be required to determine the possible direct and/or indirect effects of SARS-CoV-2 infection on upregulation of GRP78 in pneumocytes. Our observations provide new insights into stress-response pathways involving the proteostatic network implicated for host resilience and support the notion that targeting of GRP78 early in the disease process may disrupt an accelerating disease progression and might represent an alternative therapeutic strategy.

### What is GRP78?

The *HSPA5* gene encodes the 78-kilo-Dalton glucose regulated protein (GRP78), which is also known as *BiP* and acts as a central regulator of ER homeostasis and whose up-regulation is widely used as a sentinel marker for ER stress under pathologic conditions^16,17^. The Unfolded protein response (UPR) includes transcriptional activation of IRE1α (inositol-requiring enzyme 1α), PERK (pancreatic endoplasmic reticulum kinase), ATF6 (activating transcription factor 6) and X-box binding protein 1 (XBP1) to facilitate increases of UPR target genes encoding chaperone protein GRP78 in order to restore ER homeostasis^18,19^. In addition to the ER, GRP78 protein has been reported to have other compartment-specific locations such as the cell surface, the mitochondrion and nucleus. Chaperone GRP78 expression is recognized diffusely in normal lung cells, such as bronchial surface epithelial cells, chondrocytes of the bronchial cartilage, serous cells of the bronchial glands, and alveolar macrophages^18^. Our observations support elevated GRP78 expression in pneumocytes and alveolar macrophages (Figure 1).

### Unfolded protein response (UPR) in host homeostasis

In response to stressful conditions (infection, chemicals, ischemia, etc.), cells activate genetic programs which increase the synthesis of evolutionarily conserved stress proteins (Heat Shock Proteins and GRPs) as the first line of defense to restore cellular homeostasis by maintaining proper protein folding. Competition for the host’s HSP/GRP machinery creates existential threats for both host and virus, invoking complex cell fate decisions resulting in death or survival. Viral infection induces ER stress and upregulation of the unfolded protein response (UPR); furthermore the increased synthesis and translocation of multifunctional GRP78 to the cell surface has been implicated in enhanced infection in a positive feedback loop^7,20^. Major perturbations of subsequent proteostatic stress have been stymied by inadequate progress on the development of clinical tools to monitor proteostasis *in vitro* and *in vivo* especially in humans. Our findings raise the intriguing suggestion that GRP78 expression during SARS-CoV-2 infections could be an excellent target to track proteostatic dysregulation.

### Localization of GRP78 in COVID-19 lungs

Whereas SARS-CoV-2 exploits the attachment and cell fusion via the ACE2 receptor, emerging evidence implicates other host cofactors for viral entry and disease pathogenesis^21,22^. Molecular modeling, docking and structural bioinformatics that have postulated an association between molecular chaperone GRP78 and the receptor binding domain (RBD) of the spike protein of SARS-CoV-2, has given rise to the hypothesis that GRP78 facilitates virus attachment and host cell entry^8,23^. GRP78 has been recently implicated in trafficking, localization and affording for a mechanism of dynamically regulating ACE2 expression in human lung or other receptors for SARS-CoV-2 on the cell surface^14^. In addition to TMPRSS2, GRP78 protein is found *in vitro* in airway epithelial cells and confirm broad *in situ* protein expression of GRP78 in the respiratory mucosa^9^. These observations and suggestions are of particular interest considering the increased expression of GRP78 we observed in lungs of patients who died of COVID-19 (Figure 1).

### GRP78 as a biomarker of disease severity

Significantly higher serum GRP78 protein and GRP78 mRNA levels in patients with SARS-CoV-2-positive pneumonia are described compared with individuals with SARS-COV-2-negative pneumonia and healthy group^13^; however, there was no difference between SARS-CoV-2-positive pneumonia and CT-negative COVID-19 infection group^24^. A recent proteomic analysis of 144 autopsy samples from seven organs in 19 COVID-19 patients demonstrated that of 11,394 proteins in these samples, nearly half were perturbed in the COVID-19 patients compared with controls. Cathepsin L1, rather than ACE2, was upregulated in the lung from the COVID-19 patients^25^. Not surprisingly, systemic hyperinflammation and dysregulation of glucose and fatty acid metabolism were detected in multiple organs^25^. Our report adds GRP78 to the list of proteins upregulated in lung tissue of infected patients.

### Implications of ER stress and increase GRP78 expression in COVID-19 lungs

Recent reports describe markedly elevated levels of cell free DNA (cfDNA) in the serum of patients with COVID-19, the sources of which include vascular endothelial cells and lungs.^26^, and may be markers of the severity of illness. Alveolar epithelial and endothelial cells exhibit expression of hyper-phosphorylated STAT3 and immune checkpoint molecules (PD-L1 and IDO)^27^, cells which we observed to express GRP78. Exosome formation is significantly enhanced under ER stress in humans and rodents^28,29^. There are some reports that exosomes may have *protective* functions in sepsis. Circulating platelet and endothelial cell microparticles from patients with septic shock exert a protective role against vascular hypo-reactivity in rodents^30^. There is also evidence that circulating exosomes contribute to inflammatory lung injury as well as pulmonary activation or expression of GRP78. Microparticles from septic humans increased the expression of endothelial and inducible nitric oxide synthases, cyclooxygenase-2, and nuclear factor-κβ in murine tissues. Increased levels of plasma tumor necrosis factor (TNF)-α, interleukin (IL)-6, total cell counts, polymorphonuclear (PMN) leukocyte differential cell percentages and myeloperoxidase (MPO) activity in BALF, and increased wet/dry lung weight ratios and protein concentrations in BALF are reported in mice after exosome injection but not in mice treated with exosome-depleted serum^31^. A marked increase in BiP/GRP78 expression was observed in lung homogenates of mice treated with LPS^32^. In mice genetically defective in a holoenzyme that cleaves and inactivates BiP/GRP78 were protected from LPS induced lung injury. These data support upregulated expression and *proinflammatory* roles for sepsis-induced pulmonary intracellular (and not cell surface) GRP78. These reports do not identify either the cellular sources or mediators of exosomal constituents that may modify pulmonary inflammation or expression of GRP78. They are, however, consistent with our observations of enhanced expression of GRP78 in COVID-19 autopsy studies. Our studies do not permit us to address the function of increased GRP78 expression in lungs, but they provide vital new information upon which to build upon a more complete understanding of this pathway in SARS-CoV-2 pneumonia.

## Limitations to our report

First, we are cautious not to overinterpret that increased levels of viral-induced synthesis of multifunctional GRP78 in COVID-19 decedents is causally related to lung injury or a recovery response. In this study, we selected tissue samples of survivors with lung resections for GRP78 expression as controls (Table 1). Our results show substantially elevated levels of GRP78 expression in pneumocytes and lung macrophages from patients with COVID-19 relative to those of persons with other disease processes including cancer. While small numbers of patients in all groups limit conclusions based on segmented data, the ages of the COVID-19 and non-COVID-19 patients are similar. GRP78 expression is known to be increased in malignancies^33^, but our data show low GRP78 expression in pneumocytes and macrophages in tumor-adjacent ‘normal’ lung tissue. There are differences in racial distribution in COVID-19 versus non-COVID-19 control groups with more Blacks than Whites in the COVID-19 than control samples; therefore, we cannot exclude the possibility that race may modify pulmonary GRP78 expression. However, when we limited our analyses to Whites in the COVID-19 and non-COVID-19 groups, the differences in pneumocyte and macrophage GRP78 scores remained unaffected. Our semi-quantitative analyses on increased GRP78 expression of COVID-19 autopsies do not preclude dysregulation and, perhaps, proteostasis collapse *per se* as has been reported for other cellular processes such as glucose and fatty acid metabolism, immune, angiogenesis and coagulation pathways^25,34^. Plasma levels of GRP78 are not available for our patients. Nevertheless, higher serum GRP78 protein and GRP78 mRNA levels in patients with SARS-COV-2-positive pneumonia have already been described relative to a group of individuals with SARS-COV-2-negative pneumonia or a healthy group^13^.

## Data Availability

Consistent with National Institute of Health (NIH) plans for resource sharing, academic investigators will be granted access to deidentified information upon either publication and/or completion of our published and ongoing studies.

## Acknowledgements

Funding for this project to IJB was provided by the Bruce and Janine Smith Family and the Dean’s Program Development Funds, by the Department of Pathology, Medical College of Wisconsin to HR, and ERJ was supported by Merit Review BX003833. Children’s Research Institute and the Dept of Pediatrics supported PS. The authors wish to acknowledge assistance from Christina Gara and David Zimmerman in coordination of data acquisition and analysis, and Mary Rau and Mollie Patton of the Medical College of Wisconsin Tissue Bank for help with acquisition of COVID-19 and non-COVID-19 control specimens and data.

## Ethics and approval

The Institutional Review Board for Froedtert Hospital & Medical College of Wisconsin granted approval on August 19, 2020 for a waiver of HIPAA authorization requirements at 45 CFR 164 for the purpose of records review for this project. All decedent data was approved for access in accordance with 45 CFR 164.512. Project Title: *Modeling Health Outcomes using HSP Biosignatures in COVID-19 High Risk Populations*.

## Author Contribution Statement

A.P. conducted experiments, analyzed data and wrote the manuscript. E. R. J. conceived the study, provided patient materials, analyzed and interpreted data and wrote the manuscript. Y.S. provided lung pathologist expertise, reviewed images and read and edited manuscript. J.F. performed autopsies of decedents. L.G. performed experiments related to immunohistochemistry. S.L. provided materials for immunohistochemistry. R.N. provided critical comments on the manuscript. P.N. provided critical comments on the manuscript. P.S. analyzed, interpreted and wrote the manuscript. H.R. conceived the study, conducted experiments, analyzed data and wrote the manuscript. I.J.B. conceived the study, analyzed data and wrote the manuscript. All authors read and provided feedback on the study.

